# A SYSTEMATIC REVIEW ON THE EFFICACY, SIDE EFFECTS AND INTERACTIONS OF HIBISCUS SABDARIFFA ON BLOOD PRESSURE CONTROL

**DOI:** 10.1101/2025.08.01.25332612

**Authors:** SO Odunaye-Badmus, TIA Oseni, BO Akanisi, O Elemile, EA Afolabi-Obe, MT Makinde, AB Adelowo, S Yahaya-Kongoila, IJ Dozie, FM Mutseyekwa, IN Monye

## Abstract

**Background:** Control of hypertension remains a critical global health challenge, particularly in lower to middle-income nations where access is constrained. This is complicated by the side effects associated with conventional antihypertensive medications and the preference towards natural remedies. Hibiscus sabdariffa (HS) is emerging as a non-pharmacological medicinal solution given its cardio-protective and antihypertensive effects.

**Objective:** To systematically review the evidence for the efficacy, side effects and interactions of Hibiscus sabdariffa in blood pressure management among hypertensive adults.

**Methods:** A comprehensive search of major databases (PubMed, Academia, ResearchGate, AJOL and Google Scholar) was conducted, identifying randomized controlled studies, systematic reviews and review articles published from January 2015 up till September 2024 that assessed the antihypertensive effects of Hibiscus sabdariffa. The review was registered in PROSPERO (PROSPERO2025CRD420251007414) and followed the PRISMA guidelines. The study considered hypertensive adults aged 18 to 64 years and HS was tested as monotherapy. Risk of bias was evaluated using Cochrane Handbook guidelines.

**Results:** From the 884 articles retrieved, 18 studies matched the inclusion criteria and were reviewed. HS was shown to decrease both systolic and diastolic blood pressure across different ethnic groups and population centres. HS was shown to have equivalent efficacy as standard antihypertensive agents in comparative studies such as with hydrochlorothiazide and lisinopril. Additional findings suggest HS may also have lipid-lowering, antidiabetic, and organ-protective effects. Minimum adverse effects were reported and safety profile was generally favourable. The potential for herb-drug interaction with diuretics was noted but remains insufficiently explored.

**Conclusion:** *Hibiscus sabdariffa* shows promise as an effective, safe, and affordable alternative or adjunct in the management of mild to moderate hypertension. There is, however, a need for more standardized trials to establish dosage, treatment duration, and interactions with conventional antihypertensive medications.

## INTRODUCTION

Hypertension is the leading preventable cause of cardiovascular disease (CVD), resulting in mortality and morbidities from damage to the heart and blood vessels, kidneys, eyes and brain.^1^ Hypertension in adults can be defined as a persistent systolic blood pressure of equal to or greater than 140mmHg and/or diastolic blood pressure of equal to or greater than 90mmHg.^1,2,3^ Despite advances in the knowledge and management of hypertension, there has been an increase in the number of adults with hypertension globally since 1975, rising from 594 million to 1.13 billion in 2015, largely in low- and middle-income countries (LMICs).^2,3^ Due to several factors, it has been predicted that the global prevalence of hypertension is to rise to 1.5billion by the year 2025 with the rise in sub-Saharan Africa being 125.5 million in the same period.^4^ The Disability-Adjusted-Life-Years (DALY) due to hypertension is also expected to rise from 95.9million to 143.0 million between 1975 and 2015.^5^

The global view of hypertension prevalence among 182 countries stated that hypertension burden varies widely ranging from 13% to 41%.^6^ Each year, about 7.6 million deaths occur because of high blood pressure which represents 13.5% of all deaths globally.^7^ Approximately two-thirds of the world’s population of adults (aged 30-79 years) living with hypertension reside in the low-and middle-income countries.^1,8,9^

In Nigeria, according to a systematic review of cross-sectional studies done from 1995-2020, the crude prevalence of hypertension was 30.6% (95% CI: 27.3%-34.0%).^10^ When adjusted for age, study period, and sample, absolute cases of hypertension increased by 540% among adults (aged 20 years and above) from approximately 4.3 million individuals in 1995 (age-adjusted prevalence 8.6%, 95% CI: 6.5-10.7) to 27.5 million in 2020 (age-adjusted prevalence 32.5%, 95% CI: 29.8-35.3).^10^

Only one in five, that is 21%, of adults living with hypertension achieve control.^11^ This is despite the considerable increase in the types of antihypertensive medications in the past few decades.^12,13^ Aside from this, the use of antihypertensives may be associated with some adverse reactions which is likely to result in non-adherence to therapy, increased morbidity and mortality, as well as economic consequences.^13^ Adverse reactions in outpatient care have been estimated to occur in one out of four patients with hypertension.^14^ Aside from the potential side effects of these medications, the rising costs of health care and pharmaceuticals have also led to non-adherence and the search for alternative means of managing hypertension.^13,15^ Also, positive cultural beliefs and experiences among some people with hypertension often make them more willing to trust and use herbal medications.^15^

Dietary interventions, such as the use of garlic, onion, Chinese herbal teas, black and green tea, and *Hibiscus sabdariffa (HS),* otherwise known as sour team, have been shown to have some positive effects on blood pressure control.^16–20^ The use of *Hibiscus sabdariffa* as an adjunct therapy in the management of hypertension may be ideal for developing countries as it is relatively affordable, easy to grow, can be grown as part of a multi-cropping system, and can be used as food.^21^ It has also been used for medicinal purposes in China, Iran, West Africa and various other regions of the world.^16–21^ Sour tea contains carbohydrates, proteins, fatty acids, flavonoid, minerals, and vitamins. Some studies have described its anticancer, antibacterial, antioxidant, nephro- and hepato-protective, diuretic, anti-cholesterol, anti-diabetic, and anti-hypertensive properties.^21,22^ Thus its use has gained popularity among some patients with hypertension because of its perceived benefits and minimal side effects.^21,22^

Despite the frequent usage of this dietary intervention among people with hypertension, there is a dearth of evidence,^23^ especially in Africa and particularly in the Nigerian context, on its efficacy in patients with hypertension. This study aimed to bridge this gap by reviewing studies on the efficacy, potential side effects, and food-to-drug interactions of *hibiscus sabdariffa* drink on blood pressure control.

### Primary objectives

1. To evaluate the efficacy of *H. sabdariffa* in reducing systolic and diastolic blood pressure in persons with hypertension.
2. To assess the safety profile of *H. sabdariffa*, including potential side effects and adverse events.
3. To evaluate the potential interactions of *H. sabdariffa* with antihypertensive medications.

### Secondary objectives

1. To identify the potential effects of *H. sabdariffa* on other diseases.
2. To evaluate the potential interactions of *H. sabdariffa* with other drugs.

## Methodology

### Protocol and Registration

Study inclusion criteria and analysis used in the study adhered to the Preferred Reporting Items for Systematic reviews and Meta-Analyses (PRISMA). The protocol was registered with PROSPERO (PROSPERO2025CRD420251007414) on the 20^th^ of March 2025.

### Search Strategy

The database of PubMed, Academia, ResearchGate, AJOL and google scholar were systematically searched from January 2015 up to September 2024, to assess the most recent evidence. The search was performed by two independent reviewers using a combination of MeSH terms and keywords related to population, hibiscus sabdariffa, blood pressure, and blood pressure control. Some of the searched terms were: (*Hibiscus sabdariffa* [mesh terms] OR (*Hibiscus sabdariffa AND Blood pressure*) OR (*Hibiscus sabdariffa AND blood pressure control)* OR (*Hibiscus sabdariffa AND Hypertension)* OR (*Hibiscus sabdariffa AND Antihypertensives OR Antihypertensive drugs OR Hypertension medication OR Hypertension drugs*).

A total number of 884 articles were retrieved. They were screened and duplicates removed leaving 743 articles which were further screened based on the eligibility criteria to 324. The abstracts of the 324 articles were reviewed and 238 were excluded as they did not fully meet the selection criteria. The full text of 68 articles were reviewed by independent reviewers and 50 were further excluded and the remaining 18 most relevant articles were selected. This process is shown in the PRISMA chart in Figure 1.

**Figure 1:**
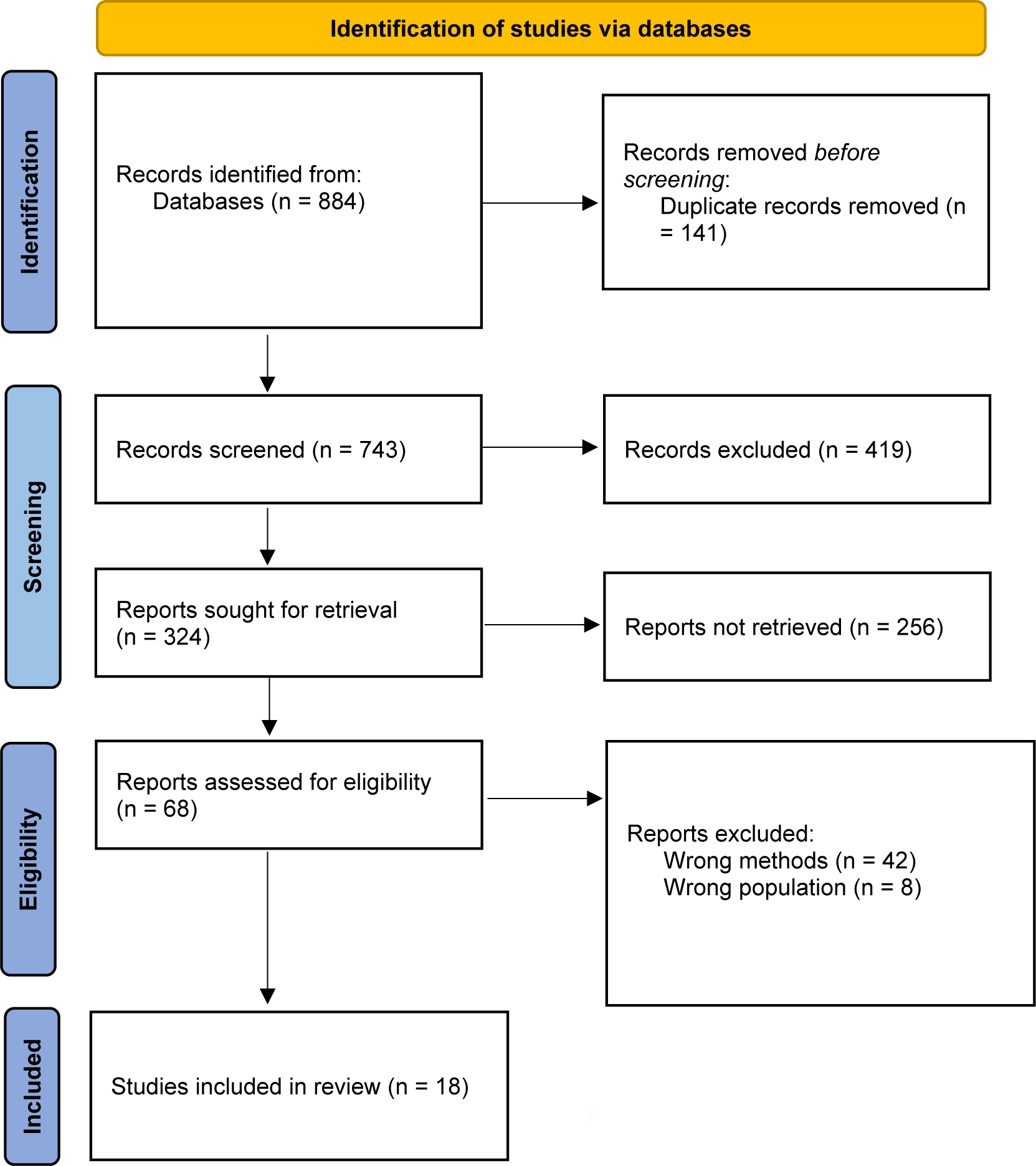
The PRISMA flow chart for the systematic review.

### Inclusion Criteria

The inclusion criteria were: (1) Papers from all regions of the world, including randomised controlled trials, and experimental studies. (2) Studies on the use of *Hibiscus Sabdariffa* in blood pressure control; (3) Studies conducted among adults aged 18 to 64 years who were hypertensive; (4) Articles published from January 2015 to September 2024; and (5) Articles written in English language.

### Exclusion criteria

The exclusion criteria were: (1) Studies involving other foods or herbs in combination with *Hibiscus Sabdariffa*; (2) Non experimental studies.

### Data Collection Process and Data Items

Two reviewers independently searched the database and all selected articles were retrieved for closer examination based on the eligibility criteria. Differences in data extraction was resolved by reference to original articles and discussion to establish consensus. The extracted data were recorded in standardized data abstraction form (Microsoft Excel®). Data points collected include title, authors, year of publication, location of study, type of study, sample size, diagnostic criteria, nature of the intervention, outcomes and other key findings, and limitations of the studies.

### Risk of Bias

All included articles were subjected to an assessment of risk of bias based on guidelines outlined by the Cochrane Handbook for Systematic Reviews of Interventions (Cochrane Collaboration, 2007). All review co-authors were provided with materials that standardize reporting of specific domains to identify potential areas of bias as part of their evaluation of study quality. The Newcastle-Ottawa scale was used to appraise the quality of the studies that were included in the study.

## Results

This study encompasses a total of 732 participants across 18 studies. Most of the studies 11 (61.1%) were randomized control trials (23, 27, 28, 31, 32, 33, 34, 38, 39, 44, and 46), 2 (11.1%) were experimental studies on humans (30, and 36), and 5 (27.8%) were experimental studies on wister rats (26, 37, 40, 41, 42) (Table 1).

**Table 1:**
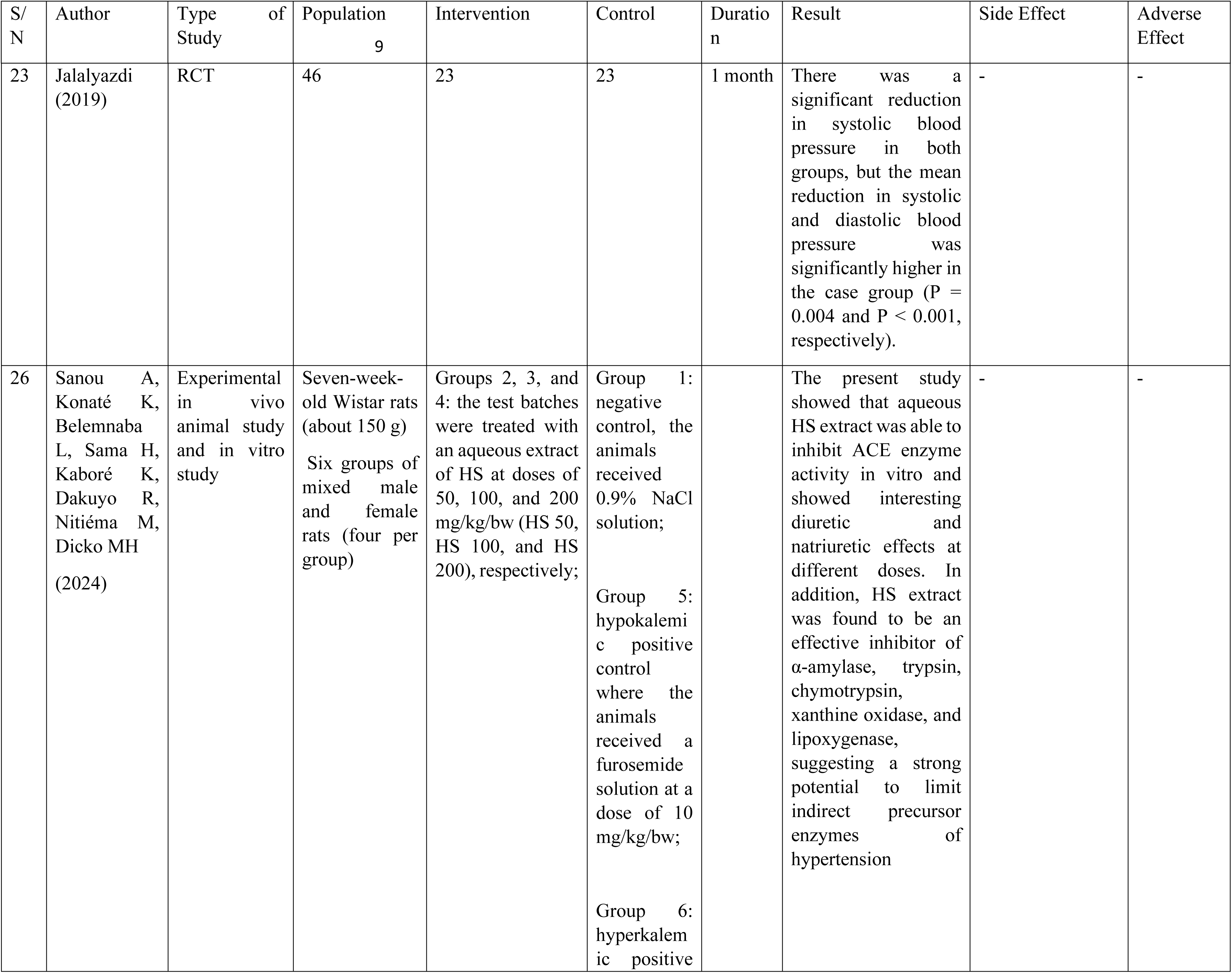
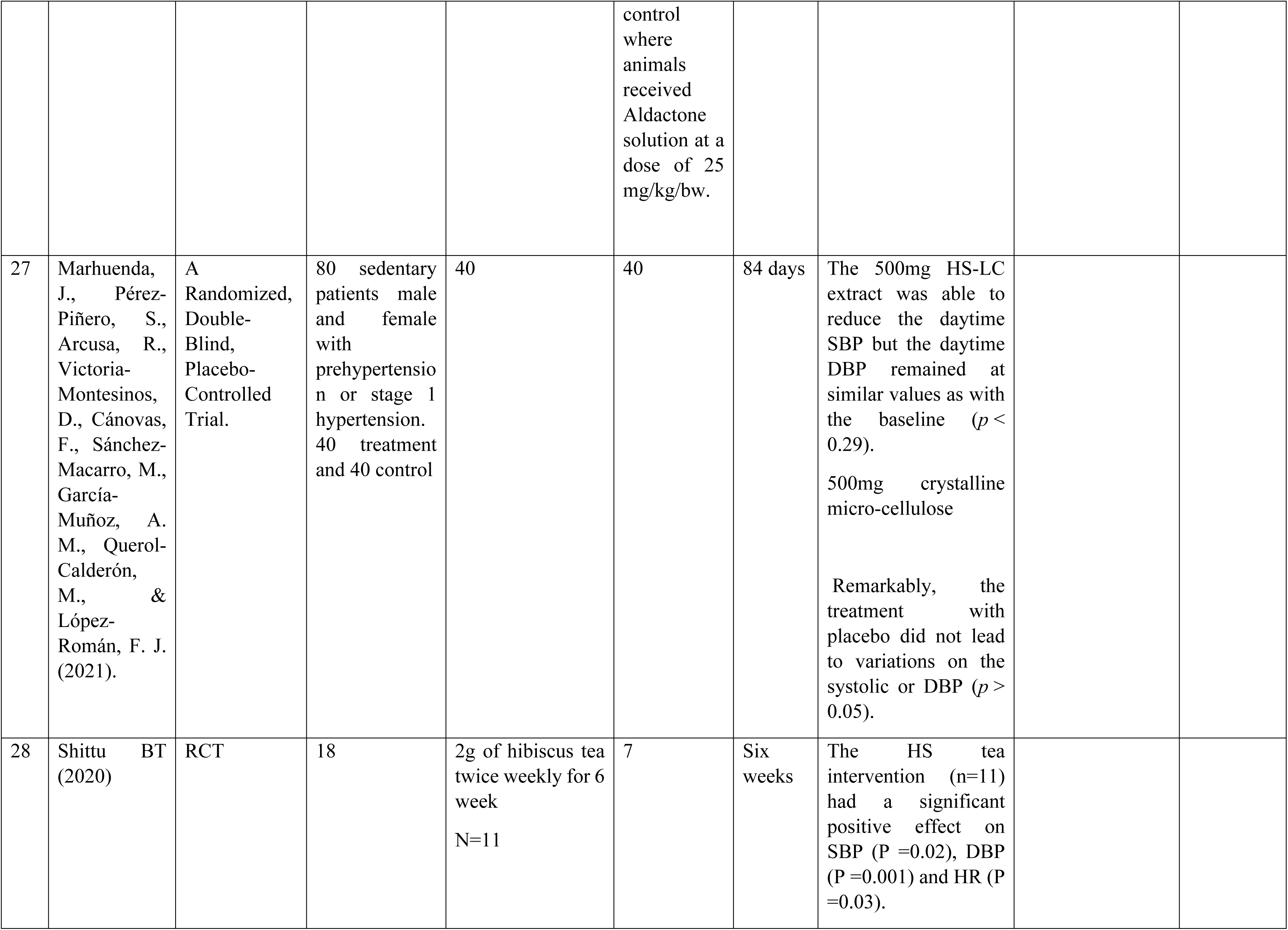
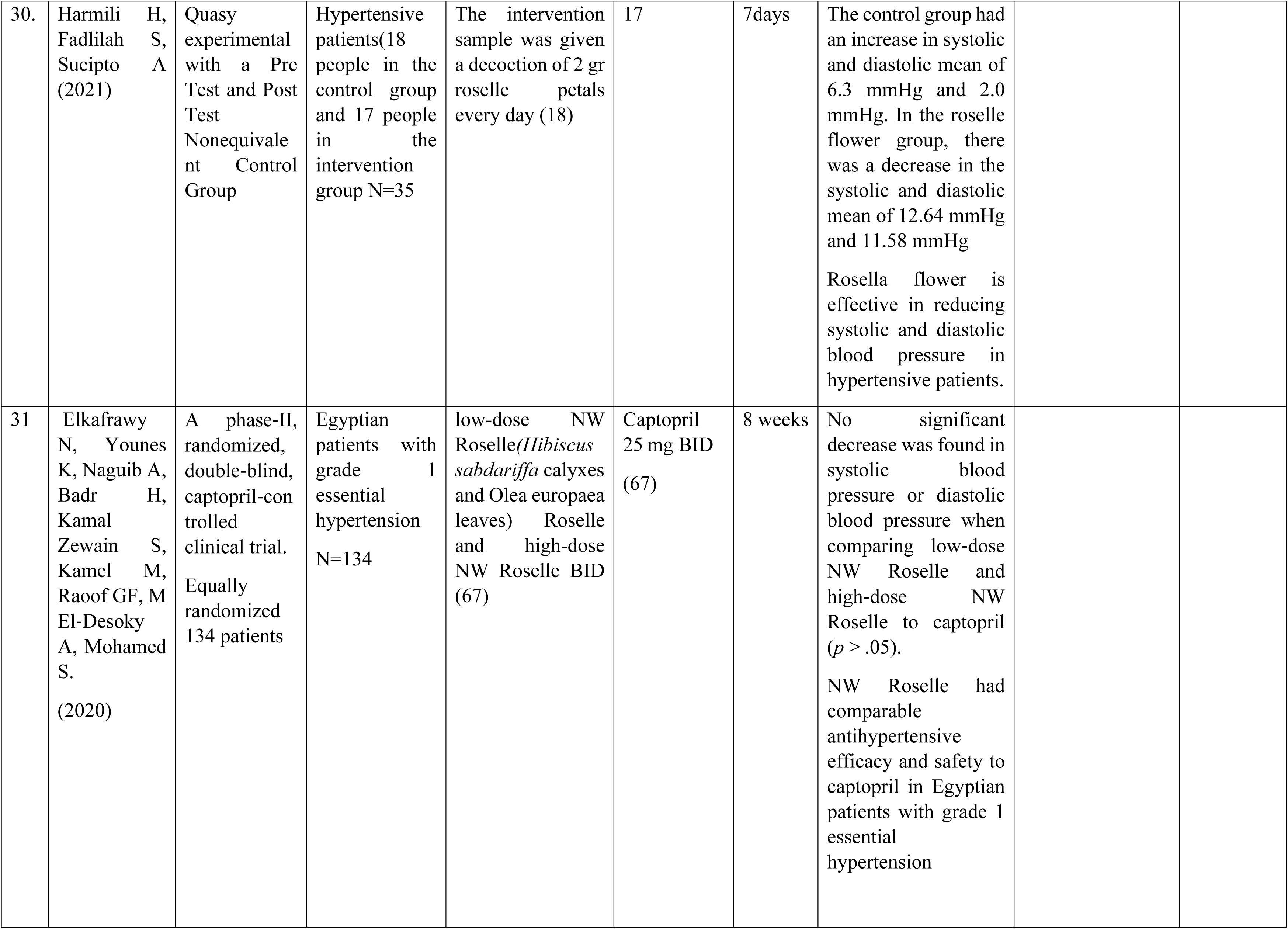
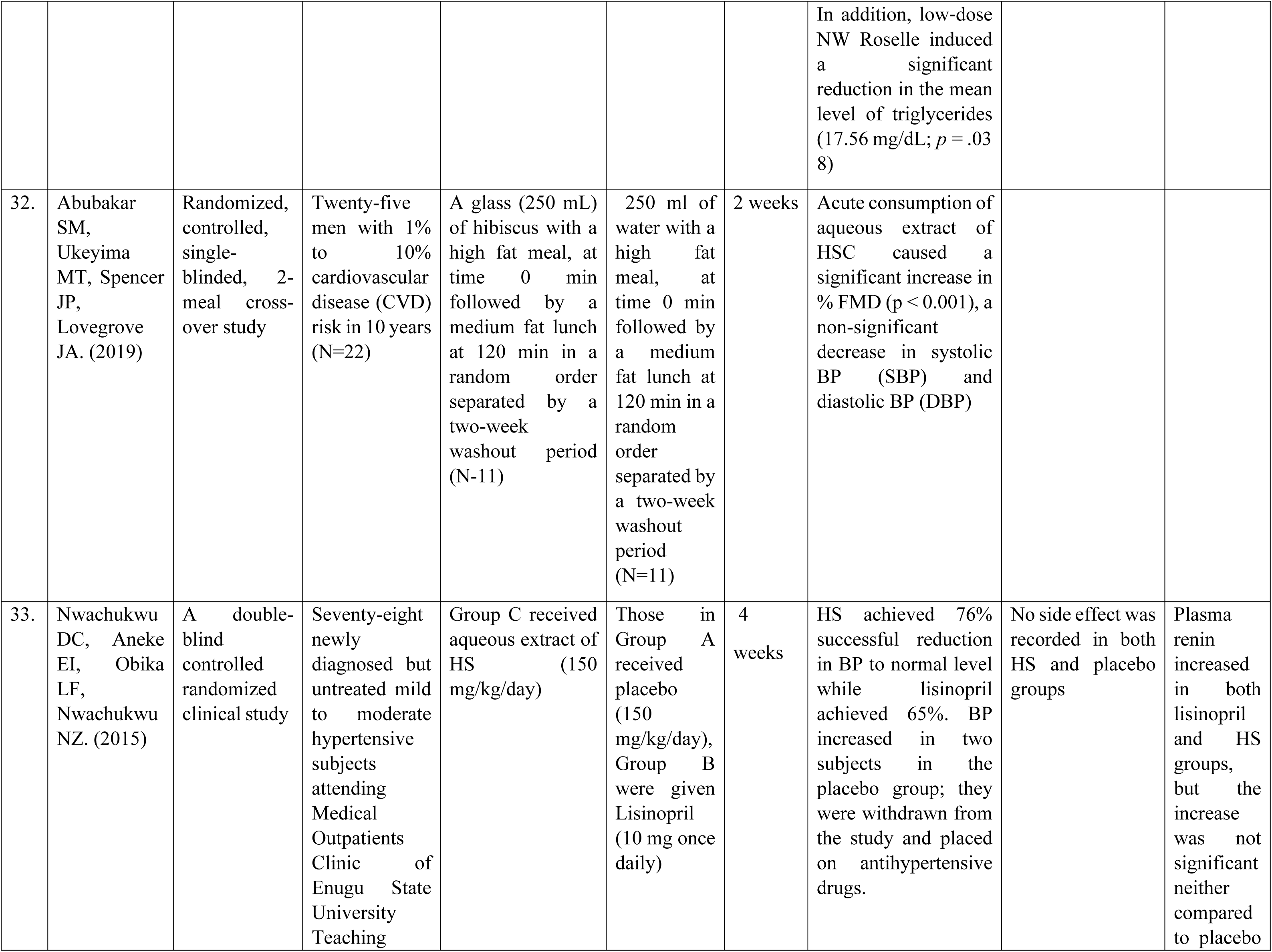
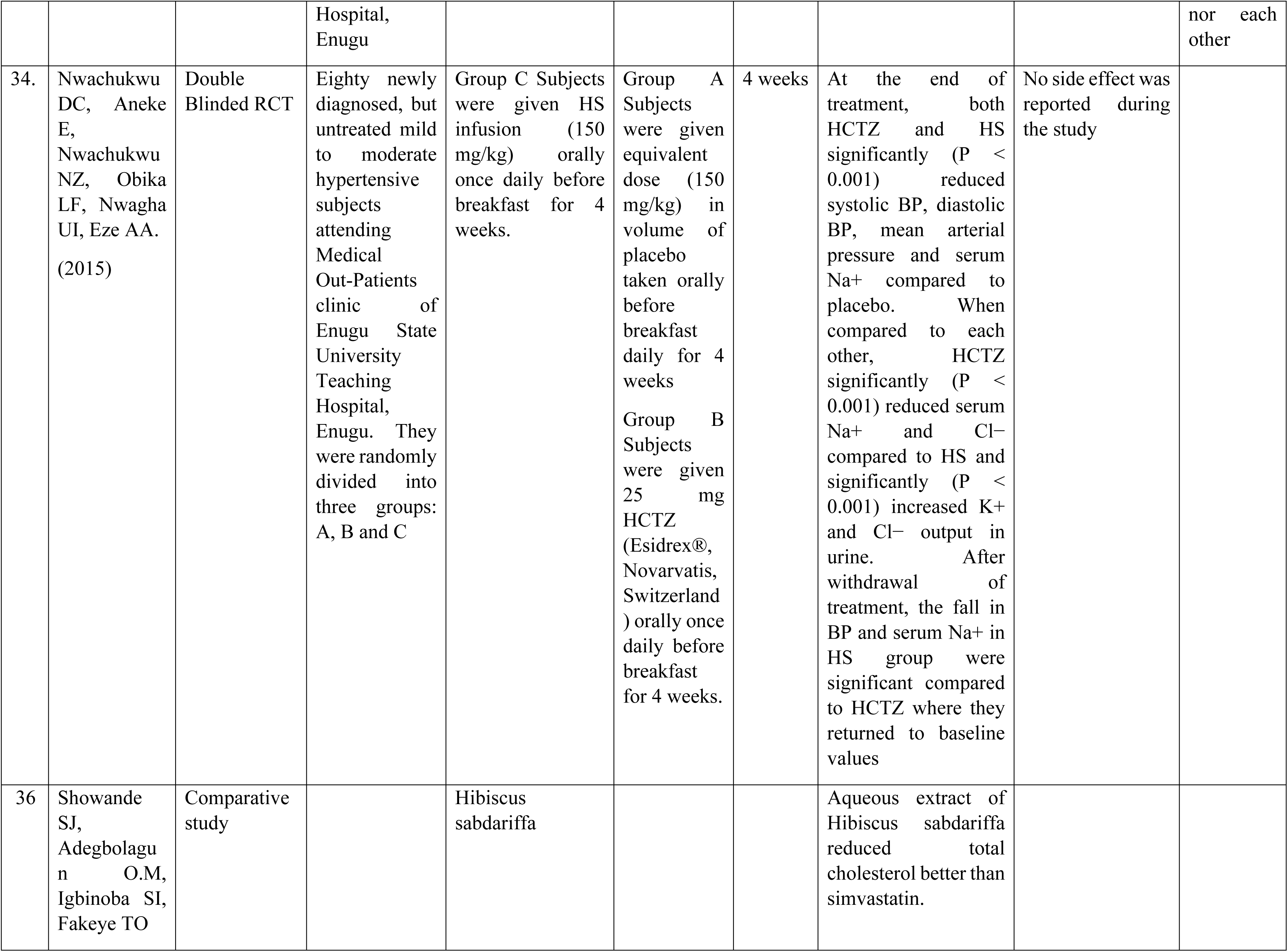
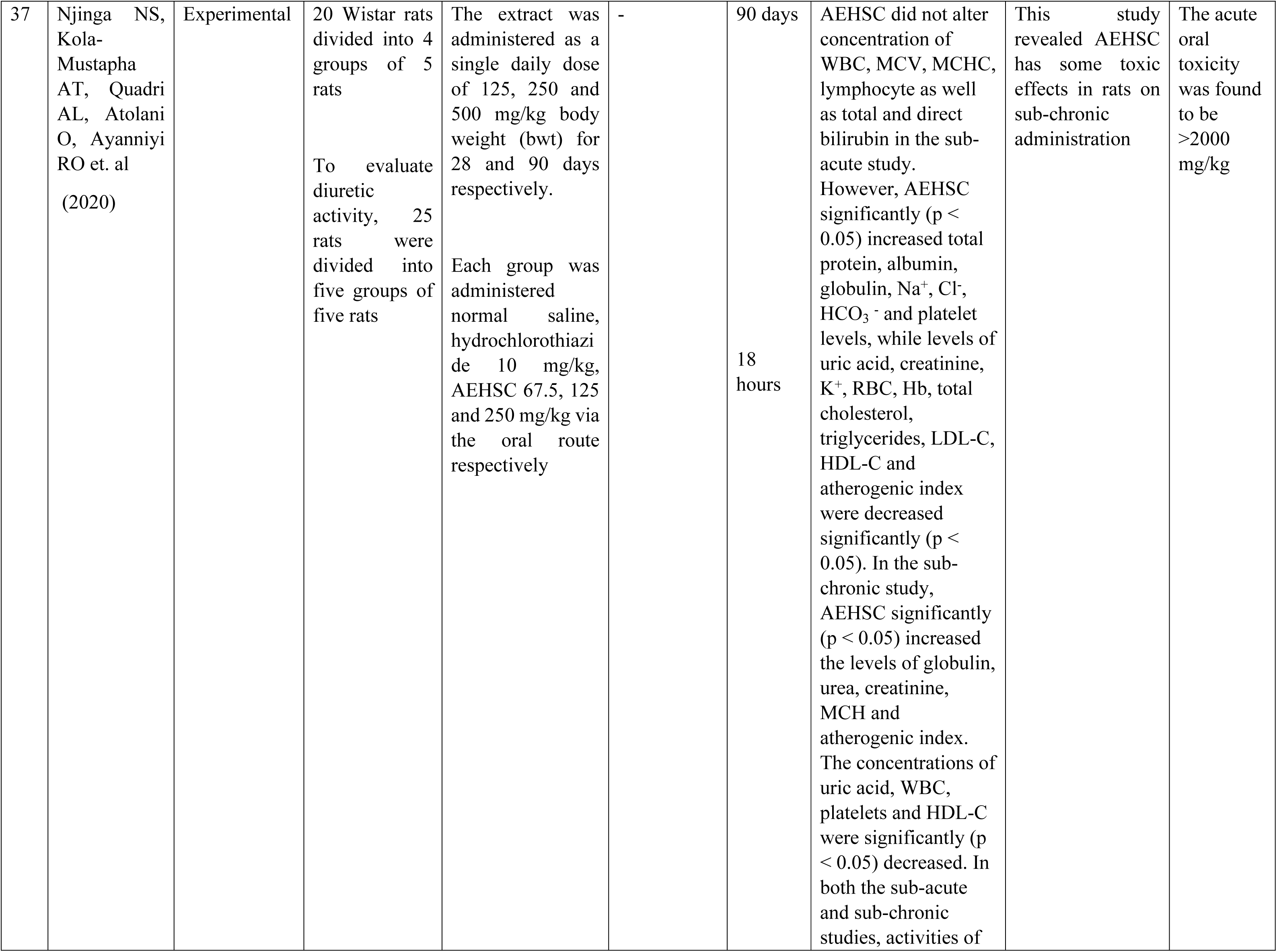
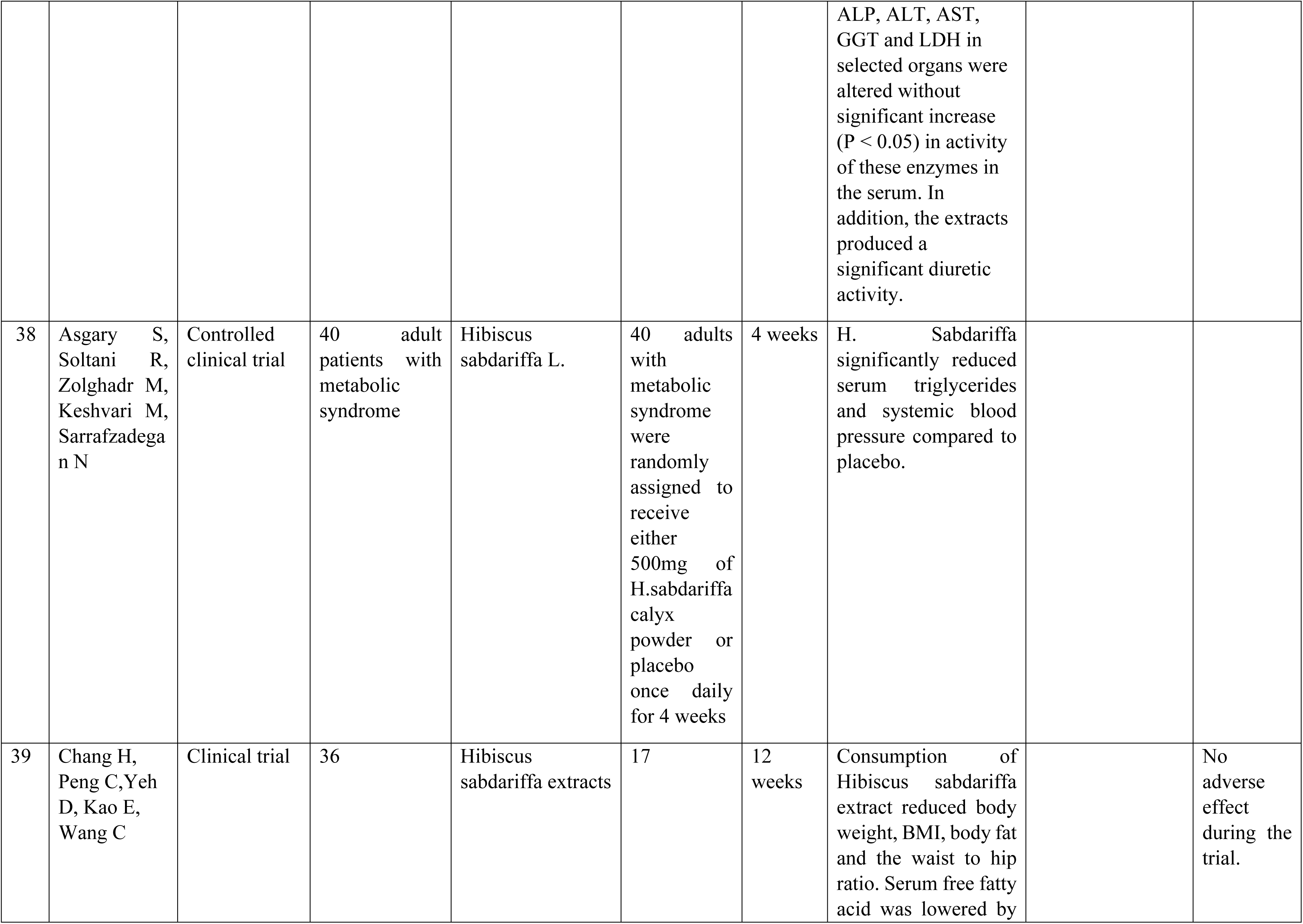
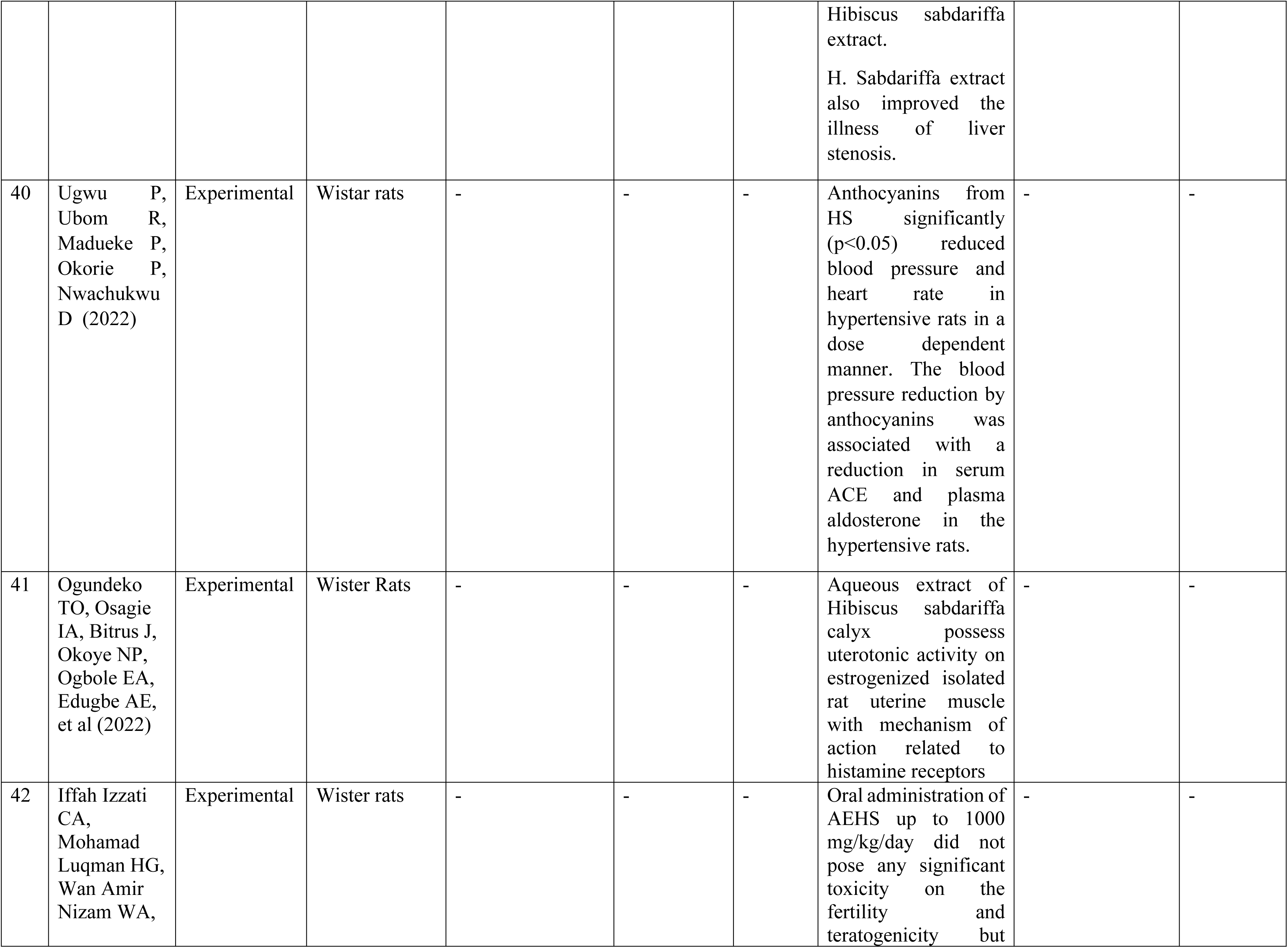
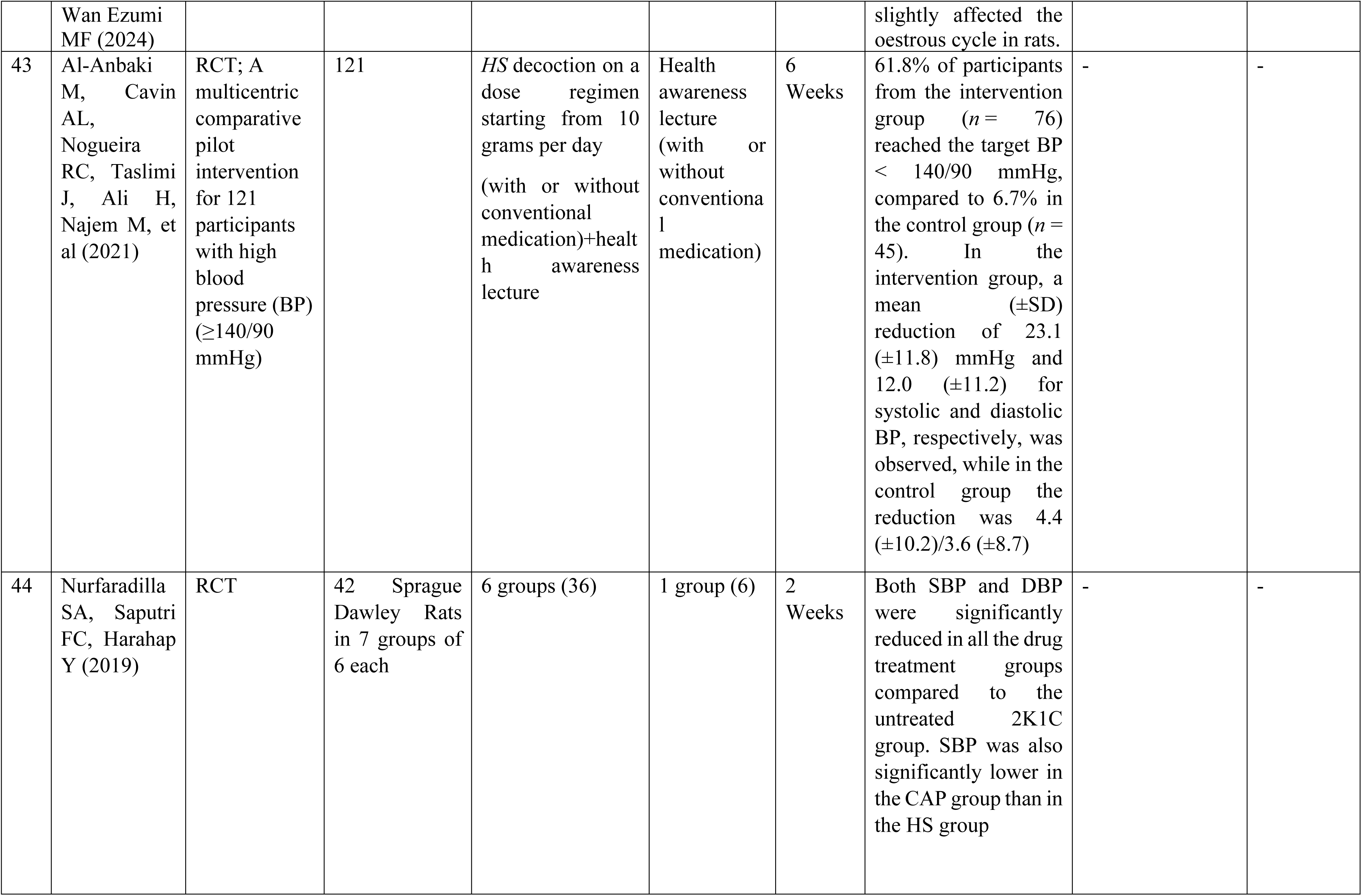
Study Characteristics.

## Discussion

*Hibiscus sabdariffa* (hibiscus), also called sour or roselle tea, is a bushy plant in the Malvaceae family.^24,25^ Studies have shown that it has multiple bioactive compounds such as carotenoids, tannins, polyphenols, and ascorbic acid.^24,25^ However, its composition often varies depending on factors such as maturity of the plant, part of the plant, the specific genotype of the plant, and climatic conditions.^24^ The calyces of hibiscus, in particular, have high concentrations of anthocyanins, mainly cyanidin 3-sambubioside and delphinidin 3-sambubioside.^24,25^ It is believed that these bioactive compounds in hibiscus have anti-inflammatory and anti-oxidant properties that may positively impact CVD biomarkers.^24,25^

The potential multiple mechanisms of actions of hibiscus on blood pressure control have been demonstrated by some animal studies.^24^ It has been proposed that the hibiscus reduce blood pressure by increasing diuresis or vasodilation/vasorelaxation. The diuretic effects are thought to be through the increase in renal filtration rate or by increasing the renal excretion of sodium and chloride. Whereas the vasodilation or vasorelaxation effect is thought to be the inhibition of calcium influx into vascular smooth muscle cells.^24,25^

Furthermore, in-vitro studies have shown that cyanidin 3-sambubioside and delphinidin 3-sambubioside can competitively inhibit the active site of the angiotensin-converting enzyme (ACE).^24^ Thus, hibiscus may be an effective competitive inhibitor of ACE, actively preventing the production of angiotensin II, with resulting reduction in blood pressure.^24,26^ Also, some in-vitro studies have shown that that delphinidin and anthocyanins cyanidin can directly inhibit renin-angiotensin system via the downregulation of inflammation and the increase in nitric oxide production by the endothelium, which may result in blood pressure reduction.^24,25^

### Effectiveness of *Hibiscus sabdariffa* in blood pressure control

An experimental study in 24 seven-week old wister rats in six groups where Group 1 was a negative control, and the animals received 0.9% NaCl solution. Groups 2, 3, and 4 test batches were treated with an aqueous extract of HS at doses of 50, 100, and 200 mg/kg/bw (HS 50, HS 100, and HS 200), respectively. Group 5 was hypokalemic positive control where the animals received a furosemide solution at a dose of 10 mg/kg/bw and Group 6 was hyperkalemic positive control where animals received Aldactone solution at a dose of 25 mg/kg/bw. The study revealed that aqueous HS extract was able to inhibit ACE enzyme activity in vitro and showed interesting diuretic and natriuretic effects at different doses. In addition, HS extract was found to be an effective inhibitor of α-amylase, trypsin, chymotrypsin, xanthine oxidase and lipoxygenase, suggesting a strong potential to limit indirect precursor enzymes of hypertension.^26^

An European study conducted in Spain, for the systolic blood pressure, the intake of the *Hibiscus sabdariffa* (HS) and *Lippia citriodora* (LC) HS-LC extract reduced the initial parameters (126.4 ± 1.8 mmHg), particularly at day 56 (−3.76 ± 10.13 mmHg; *p* < 0.05). This was not observed for the placebo, which showed similar values throughout the whole study (*p* > 0.05). The comparison between the placebo and HS-LC groups along the whole study led to significant differences in the SBP (*p* < 0.02). Therefore, the HS-LC extract appeared to improve the SBP to a greater extent compared with the placebo, which was observed from the first 56 days of treatment. The diastolic blood pressure was however unchanged significantly compared to the placebo group in this study (p=0.19).^27^

In an open Master’s thesis from the University of United States, Eighteen participants completed a study by consuming 2g of Hibiscus sabdariffa or oolong tea twice daily for six weeks. Blood pressure, pulse rat and Central arterial stiffness was analyzed as cfPWV using applanation tonometry technique. The HS tea intervention (n=11) had a significant positive effect on SBP (P =0.02), DBP (P =0.001) and HR (P =0.03). Also, the HS tea consumption led to a non-significant (p=0.44) reduction in cfPWV (−0.5m/ s) when compared to control tea (+0.3m/s).^28^

A randomized controlled clinical trial was conducted to evaluate the effectiveness of sour tea (*H. sabdariffa*) on patients with stage one hypertension referred to an outpatient cardiology clinic in Imam Reza Hospital in Mashhad, Iran. Every patient who was aged between 18 and 70 years with stage 1 hypertension. These patients had systolic and diastolic blood pressure ranging from 130 to 139 and 80–89 mmHg, respectively.^23^ A total of 46 participants (23, 50% in the tea group and 23, 50% in control group) participated in this study.^23^ Twenty-three patients in experimental group received nonmedical treatment advices and two standard cup of sour tea (each cup with one sour tea bag containing 1.25 g *H. sabdariffa* (480 m .L/d)) every day morning and night for 1 month (they did not use any other kind of teas).^23^ The other group only had dietary counselling by a nutrition specialist; decreasing sodium and increasing potassium under the consideration of a nutrition specialist, and doing aerobic exercises at list 5 days a week for 30 min.^23^ The blood pressure after the study period showed significant reduction in systolic blood pressure in both groups (*P* < 0.05), but the mean reduction in systolic blood pressure was significantly higher in the intervention group (−7.43 mmHg) compared to the control group (−1.91 mmHg) (*P* = 0.004).^23^ There was a significant reduction in diastolic blood pressure in both groups (*P* < 0.05) but the mean reduction in diastolic blood pressure was significantly higher in the intervention group (−6.70 mmHg) compared to the control group (−3.96 mmHg) (*P* < 0.001).^23^ The main limitation of suggesting *H. sabdariffa* as a blood pressure lowering agent or for other cardiovascular risk reductions is the heterogeneity of clinical trials’ protocols because different therapeutic doses has been reported for achieving the beneficial effect of sour tea.^23^

In Indonesia a quasy experiment with a Pre Test and Post Test Nonequivalent Control Group was conducted with 18 people in the control group and 17 people in the intervention group.^30^ The intervention sample was given a decoction of 2 gram Roselle petals every day for seven days.^30^ The control group had an increase in systolic and diastolic mean of 6.3 mmHg and 2.0 mmHg while in the Roselle flower group, there was a decrease in the systolic and diastolic mean of 12.64 mmHg and 11.58 mmHg.^30^ The difference between the changes in the mean posttest-pretest systolic and diastolic control and Roselle groups was 18.94 mmHg and 13.58 mmHg.^30^ The bivariate results of the systolic and diastolic pretest-posttest in the Roselle flower group obtained p-values < of 0.000 and 0.000.^30^ Comparison of systolic and diastolic blood pressure in the control group and rosella flowers obtained p-values < of 0.000 and 0.000. Rosella flower is effective in reducing systolic and diastolic blood pressure in hypertensive patients.^30^

The antihypertensive efficacy and safety of two doses of an herbal product of Hibiscus sabdariffa calyxes and Olea europaea leaves (NW Roselle) in Egyptian patients with grade 1 essential hypertension was conducted.^31^ A total of 134 patients were equally randomized to receive captopril 25 mg, low-dose NW Roselle, or high-dose NW Roselle BID for 8 weeks.^31^ No significant decrease was found in systolic blood pressure or diastolic blood pressure when we compared low-dose NW Roselle and high-dose NW Roselle to captopril (p > .05).^31^ In all groups, mean reduction in BP at 8 weeks was significant; 16.4/9.9 mmHg (p < .0001), 15.4/9.6 mmHg (p < .0001), and 14.9/9.4 mmHg (p < .0001) with captopril, low-dose NW Roselle, and high-dose NW Roselle respectively.^31^ In addition, low-dose NW Roselle induced a significant reduction in the mean level of triglycerides (17.56 mg/dl; p = .038).^31^ It was concluded that NW Roselle had comparable antihypertensive efficacy and safety to captopril in Egyptian patients with grade 1 essential hypertension.^31^

In a study on the acute effect of H. Sabdariffa on blood pressure change in SBP, DBP, PP and heart rate up to 4 h after hibiscus drink or water consumption relative to baseline showed no significant change.^32^ There was no significant effect of treatment, time or treatment-time interaction for SBP or DBP up to 4 h post hibiscus drink consumption.^32^ Significant (*p* = 0.003) time effect was observed for the heart rate and PP (*p* = 0.011) while no significant effect of treatment or treatment-time interaction was observed. No significant difference was observed in any of the summary response measure when hibiscus was compared to water.^32^

Nwachukwu and colleagues in a double-blind controlled randomized clinical study was used with seventy-eight newly diagnosed but untreated mild to moderate hypertensive subjects attending Medical Outpatients Clinic of Enugu State University Teaching Hospital, Enugu were recruited.^33^ The study investigated the effects of aqueous extract of Hibiscus sabdariffa (HS) on the three basic components of renin-angiotensin-aldosterone system: Plasma renin, serum angiotensin-converting enzyme (ACE), and plasma aldosterone (PA) in mild to moderate essential hypertensive Nigerians and compared with that of Lisinopril, an ACE inhibitor.^33^ Those in Group A received placebo (150 mg/kg/day), Group B were given Lisinopril (10 mg once daily) while those in Group C received aqueous extract of HS (150 mg/kg/day).^33^ After 4 weeks of treatment, the levels of plasma renin, serum ACE, and PA were determined which showed that HS and Lisinopril significantly (P < 0.001) reduced PA compared to placebo by 32.06% and 30.01%, respectively.^33^ Their effects on serum ACE and plasma renin activity (PRA) were not significant compared to placebo; they reduced ACE by 6.63% and 5.67% but increased plasma PRA by 2.77% and 5.36%, respectively.^33^ This study concluded that HS reduced serum ACE and PA in mild to moderate hypertensive Nigerians with equal efficacy as Lisinopril and that these actions are possibly due to the presence of anthocyanin in the extract.^33^

In another comparative study by Nwachukwu and colleagues also in Nigeria, with participants were randomly divided into three groups and given placebo, Hydrochlorothiazide and *Hibiscus sabdariffa* treatment which lasted for 4 weeks.^34^ Blood pressure, serum and urine electrolytes were measured at baseline, weekly during treatment and a week after withdrawal of treatment.^34^ It concluded that HS was a more effective antihypertensive agent than HCTZ in mild to moderate hypertensive Nigerians and did not cause electrolyte imbalance.^34^

### Potential Effects of Hibiscus Sabdariffa on other Diseases

Based on available data, the polyphenols and anthocyanins found in roselle calyces appear to have a variety of biological effects. Numerous studies have emphasized the dried calyces of hibiscus as a possible source of bioactive molecules that have potent anti-inflammatory, antioxidant, antiradical, antihypertensive, anti-obesity, antihyperlipidemic, diuretic, anti-agglutination, antiurolithicatic, anti-cancer, anti-microbial, reno-protective, and hepatoprotective properties.^35^

A comparative study that investigated the effects of aqueous beverage of Hibiscus sabdariffa (ABHS 250 mg/kg) and simvastatin (10mg/kg) in hyperlipidemia-induced Wistar rats for 2 weeks noticed that the ABHS was more effective in total cholesterol reduction, compared to simvastatin (P = .031).^36^ A study conducted on the toxicity assessment of sub-acute and sub-chronic oral administration and diuretic potential of AEHSC in Wister rats reported that AEHSC did not alter concentration of WBC, MCV, MCHC, lymphocyte as well as total and direct bilirubin in the sub-acute study. However, AEHSC significantly (p < 0.05) increased total protein, albumin, globulin, Na+, Cl-, HCO3 - and platelet levels, while levels of uric acid, creatinine, K+, RBC, Hb, total cholesterol, triglycerides, LDL-C, HDL-C and atherogenic index were decreased significantly (p < 0.05). In the sub-chronic study, AEHSC significantly (p < 0.05) increased the levels of globulin, urea, creatinine, MCH and atherogenic index. The concentrations of uric acid, WBC, platelets and HDL-C were significantly (p < 0.05) decreased. In both the sub-acute and sub-chronic studies, activities of ALP, ALT, AST, GGT and LDH in selected organs were altered without significant increase (P < 0.05) in activity of these enzymes in the serum.^37^ A randomised controlled trial among 40 adults with metabolic syndrome revealed that H. Sabdariffa significantly reduced serum triglycerides.^38^ A clinical trial among 36 patients with BMI greater than or equal to 27 to confirm the effect of Hibiscus sabdariffa extract on obesity and fat accumulation revealed that consumption of Hibiscus sabdariffa extract reduced body weight, BMI, body fat and the waist to hip ratio.^39^ Serum free fatty acid was also lowered by Hibiscus sabdariffa extract and the extract also improved the illness of liver stenosis.^39^

### Safety Profile of Hibiscus Sabdariffa

A double-blind RCT among Seventy-eight newly diagnosed but untreated mild to moderate hypertensive subjects attending Medical Outpatients Clinic of Enugu State University Teaching Hospital, Enugu reported no side effect in both groups^33,34^ however plasma renin increased in both Lisinopril and HS groups, but the increase was not significant neither compared to placebo nor each other.^33^

*Hibiscus sabdariffa* is becoming more globally relevant and available in food and herbal solutions with no known record of significant harmful effects.^37^ None of these studies noticed a significant side effects or adverse events after the intake of Hibiscus. One of the included RCT studies investigated the potential side effects/adverse events after 80 Nigerian with untreated mild to moderate hypertension were given oral Hibiscus (150 mg/kg), daily for 4 weeks. The study noticed significant reduction in serum sodium with no significant electrolyte imbalance after the intake of Hibiscus.^34^ An experimental study among 20 wister rats to assess the toxicity of sub-acute and sub-chronic oral administration and diuretic potential of aqueous extract of Hibiscus sabdariffa calyces (AEHSC) reported that AEHSC had some toxic effects in rats on sub-chronic administration.^37^ The study also reported that the acute oral toxicity was found to be >2000 mg/kg.^37^ A clinical trial among 36 patients with BMI greater than or equal to 27 to confirm the effect of Hibiscus sabdariffa extract on obesity and fat accumulation reported no adverse effect during the trial.^39^

### The Uterotonic and Antihypertensive Effects of Hibiscus sabdariffa (Zobo): Implications in Pregnancy

In Nigeria, Hibiscus sabdariffa popularly known as Zobo, is frequently consumed. In recent times, it has become known for its antihypertensive benefits which are linked to phytochemicals such as anthocyanins, flavonoids, and polyphenols. These bioactive components are known to restrain angiotensin converting enzyme (ACE) causing vasodilation which leads to decreased blood pressure.^40^ While it is certainly appealing as one of the natural remedies for hypertension, its safety in pregnancy should be considered.

Several plants have been screened for their uterotonic activity using in vitro methods with positive results and report from a community visit showed that adolescent girls orally consume high concentration of Hibiscus sabdariffa calyces drink for abortion.^41^

A study on the uterotonic screening of calyx extracts of Hibiscus sabdariffa on estrogenized isolated Uterus of Wister strain albino rats reported that Aqueous extract of Hibiscus sabdariffa calyx possess uterotonic activity on estrogenized isolated rat uterine muscle with mechanism of action related to histamine receptors and contraction of the myometrial cells as the was antagonized by promethazine and Nifedipine.^41^ In the study, extracts of Hibiscus sabdariffa was dissolved in hot water, ethanol, methanol, acetone and cold water. Administration of increasing doses of hot water (2 x 10-3 g/ml), ethanol (2 x10-3 g/ml), methanol (2 x10-3 g/ml), and acetone (2 x 10-3 g/ml) produced no contraction on the uterine smooth muscle and may be due to the fact that the active constituents in Hibiscus sabdariffa are more soluble in cold water. However, the cold-water extract has higher percentage yield (15.3%) and uterotonic activity at concentration of 2 x10-3 g/ml on the isolated tissue.^41^

The study further recommended that the mode of preparation before consumption of the plant product should be looked into, women in their third trimester should apply caution in taking beverages especially such prepared via soaking H. sabdariffa cold water. There is a need to undertake further studies to assess if the same results can be seen in vivo when the herb is ingested as water, spirit, or oil-based medicines.^41^

A study on the fertility and teratogenic effect of the aqueous extract of Hibiscus sabdariffa L. in female Sprague Dawley rats. AESH treatment was administered orally by gavage at four different dosages: 250, 500 and 1000 mg/kg/day or distilled water (control). Results obtained revealed that the mean length of the oestrous cycle was not statistically affected, even though a few rats displayed irregular cycles. The study concluded that the oral administration of AEHS up to 1000 mg/kg/day did not pose any significant toxicity on the fertility and teratogenicity but slightly affected the oestrous cycle in rats.^42^

### Interactions of *Hibiscus Sabdariffa* with antihypertensive medications

The potential for hibiscus-drug interactions is another possibility with the intake of hibiscus. Although no known study has investigated the potential interactions between hibiscus and different classes of antihypertensive, the assumption is that no significant interaction is expected. This is because hibiscus is a commonly taken beverage in many communities and no known study has recorded any significant interaction between it and any antihypertensive.^43^

None of the studies included in this review also reported any interaction between hibiscus and any antihypertensive. However, in a recent study on rats, the joint intake of hydrochlorothiazide (10 mg/kg) and hibiscus (20–40 mg/kg) significantly raised urine volume over a 24-hour period, thus increasing the possibility of dehydration.^37^ Furthermore, since one of the potential mechanisms of action of hibiscus is ACE inhibition, its possible interacts or potentiating effect with ACE inhibitor medications needs to be carefully investigated and established.^37^

However, a recent study that investigated the effects of the co-administration of Hibiscus sabdariffa aqueous extract and captopril (an ACE inhibitor) in rats noticed that the Hibiscus extract did not enhance the blood lowering effects of captopril.^44^ However, more studies, especially in humans, need to be done on the potential interaction of hibiscus with ACE inhibitors and other antihypertensive medications.^37^

### Interactions of Hibiscus Sabdariffa with other medications and plant extracts

In recent times, there has been a rising interest on the subject of possible toxicity and herb-drug interactions worldwide as regards herbal medicines. Although, Hibiscus sabdariffa is utilized to cure various diseases, and it is often efficacious, its safety profile has not garnered sufficient assessment by health practitioners or patients, as proven by heightened safety concerns on the risk of toxicity and interactions with drugs, hence additional investigations are warranted on the safety of hibiscus–drug interactions.^24,45^ Similar, to the recommendations of a study on the systematic review of clinical trials on the efficacy of H.Sabdariffa in essential hypertension, stating further research should focus on dose and duration of treatment, interactions with medications and quality of starting material.^29^

However, the findings of a recent systematic review and meta-analysis (involving 7 RCTs and 332 participants) noticed that the concurrent use of Hibiscus Sabdariffa (Hibiscus) with many other plant extracts can potentiate the anti-hypertensive, anti-lipid, anti-adiposity, and blood glucose lowering effects of hibiscus.^46^ A situation that has the potential of resulting in the over control of these conditions.

Furthermore, another experimental study evaluated the effects of the combination of low dose aqueous beverage of Hibiscus sabdariffa (ABHS 250 mg/kg) and low dose simvastatin (10mg/kg) in hyperlipidemia-induced Wistar rats. The study noticed that the combined intake of low-dose ABHS and low-dose simvastatin resulted in better reduction of Total cholesterol (38.3%) and Triglycerides (57.4%) compared to the use of low-dose simvastatin.^36^ However, there was a significant herb-drug interaction, with ABHS reducing the peak concentration and increased the clearance of simvastatin by 18.0% and 44.6% (P < .05), respectively. The study concluded that the combined administration of simvastatin and Hibiscus should not be encouraged until more clinical studies are done.^36^

It is advised that health professionals must counsel their patients about potential interactions of hibiscus with other medications and plant extracts.^45^ In addition, more well-designed controlled trials in humans on the pharmacokinetics of hibiscus extracts and its various components are needed, especially on the effects of its long-term intake in humans.^45^

### Limitations

1. Lack of uniformity across studies: this stemmed from the study design, sample population, intervention dose, and intervention duration which made it impossible to undertake a meta-analysis.
2. Absence of African Focused Evidence: despite the significance of HS in African traditional medicine, there were very few good-quality randomized controlled trials (RCTs) conducted on African patients found.
3. Linguistic and Publication Bias: The review excluded non-English studies which could have contained pertinent information because only studies published in English were considered.
4. Short Intervention Period: A number of studies had short follow-up periods which may have hampered measurements of the longitudinal effectiveness and safety of the interventions.
5. Insufficient Documentation of Herb-Antihypertensive Drug Interactions: Very few studies actively explored or documented HS’s pharmacokinetic or pharmacodynamic interactions with conventional antihypertensive medications.
6. Some of the studies reviewed did not show the year it was done, the age range of participants, and the exact dosing of the hibiscus tea.
7. Also, there was no standardization in terms of the concentration of the aqueous extract / decoction and the Hibiscus tea, for better comparisons among some of the studies.

## Conclusion

This systematic review strongly suggests that Hibiscus sabdariffa is an effective natural treatment for hypertension in populations with limited access to other medical resources or where treatment is not followed due to negative side effects. Given its safety and other cardiometabolic advantages, HS has potential as a cost-effective hypertension management solution. Nevertheless, thorough investigations are essential to inform the effective development of clinical guidelines and public health strategies.

As for H. Sabdariffa and ACE inhibitors, while it does not appear to negatively influence their action, it may not provide additional antihypertensive benefit when used together with them either. H. Sabdariffa appears to be very safe, except for minor gastrointestinal disturbances and the risk of dehydration, especially when used with ACE inhibitors.

### Recommendations

1. Standardized Procedures for Interventions: Further research should employ HS dosage, preparation methods, and treatment length to enable evidence comparison and synthesis.
2. Encourage Local Clinical Trials: High-quality, large-scale RCTs incorporating local dietary habits and health beliefs should be conducted in African populations.
3. Assess Interactions Between Herbs and Drugs: Further pharmacological safety evaluation of HS alongside prescribed antihypertensives needs to be undertaken.
4. Develop Clinical Guidelines: National guidelines for managing hypertension in low-resource settings should consider HS as a supporting therapy alongside emerging best evidence.
5. Spread Awareness and Educate: Healthcare practitioners need to be educated on the proper evidence-based use of herbal therapy HS.
6. More studies are needed to establish effective dose response and treatment duration regarding the effect of H.Sabdariffa decoction taken simultaneously with each class of antihypertensive agent

## Data Availability

All relevant data are within the manuscript and its Supporting Information files

## PRISMA CHECKLIST

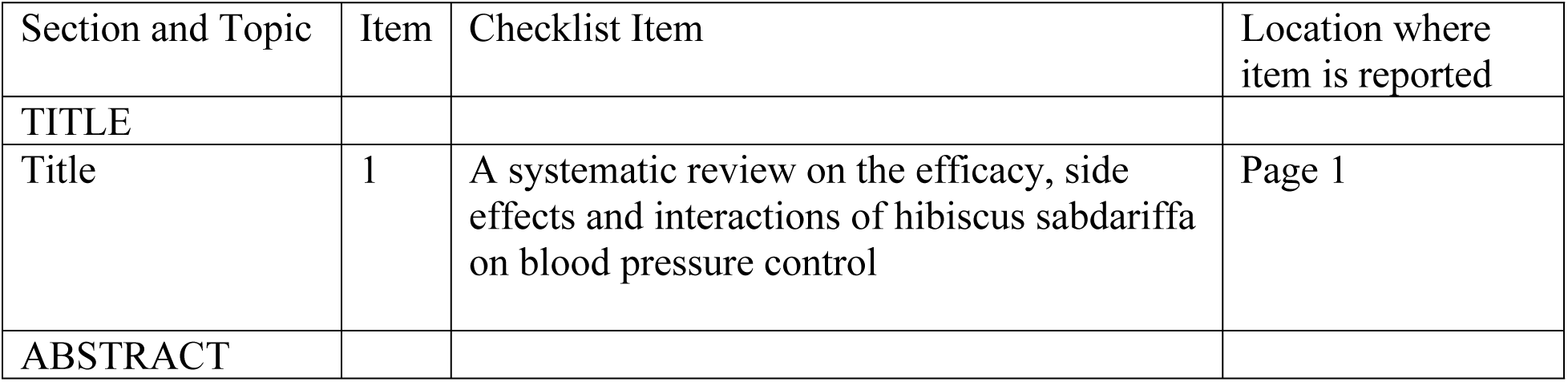
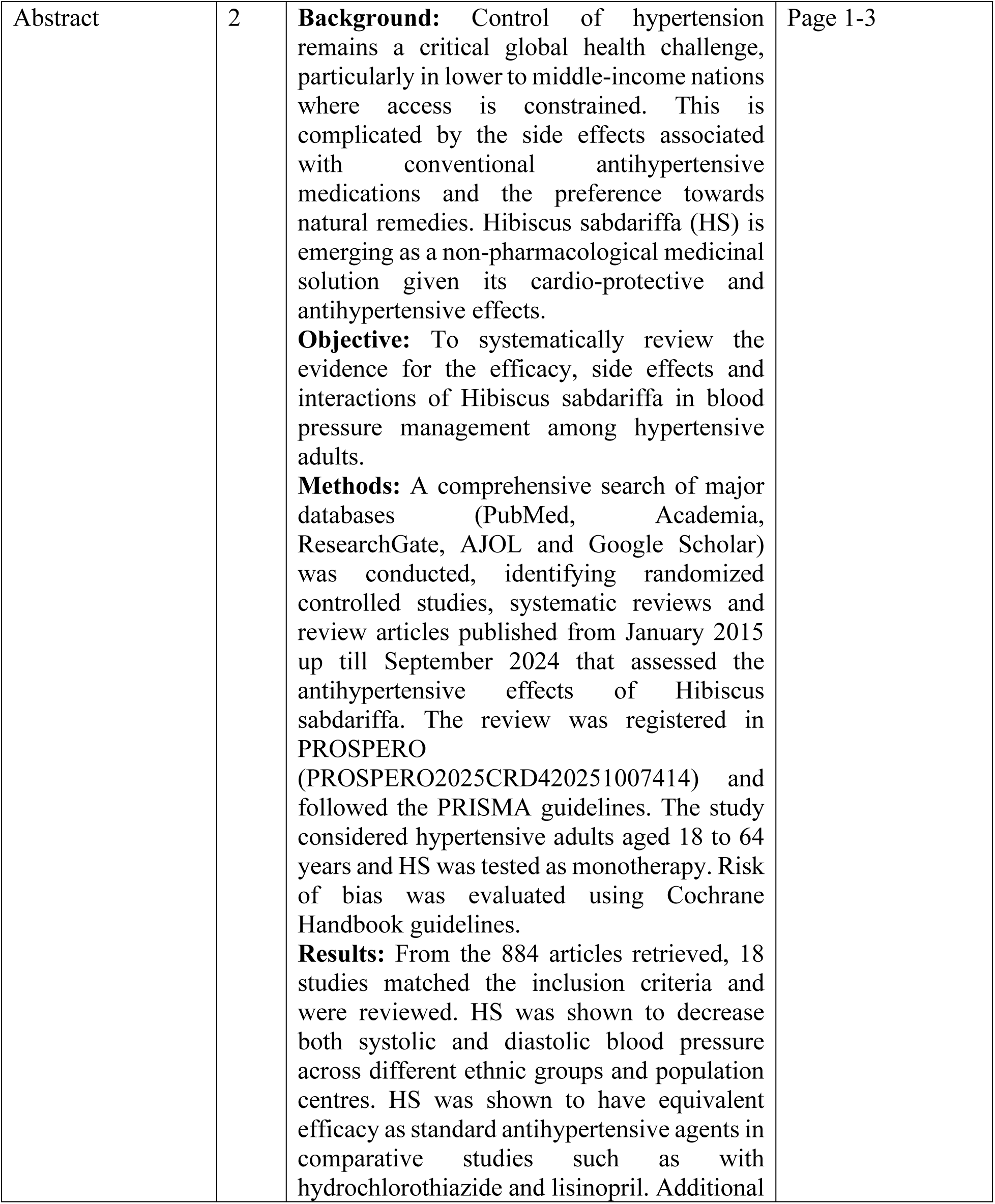
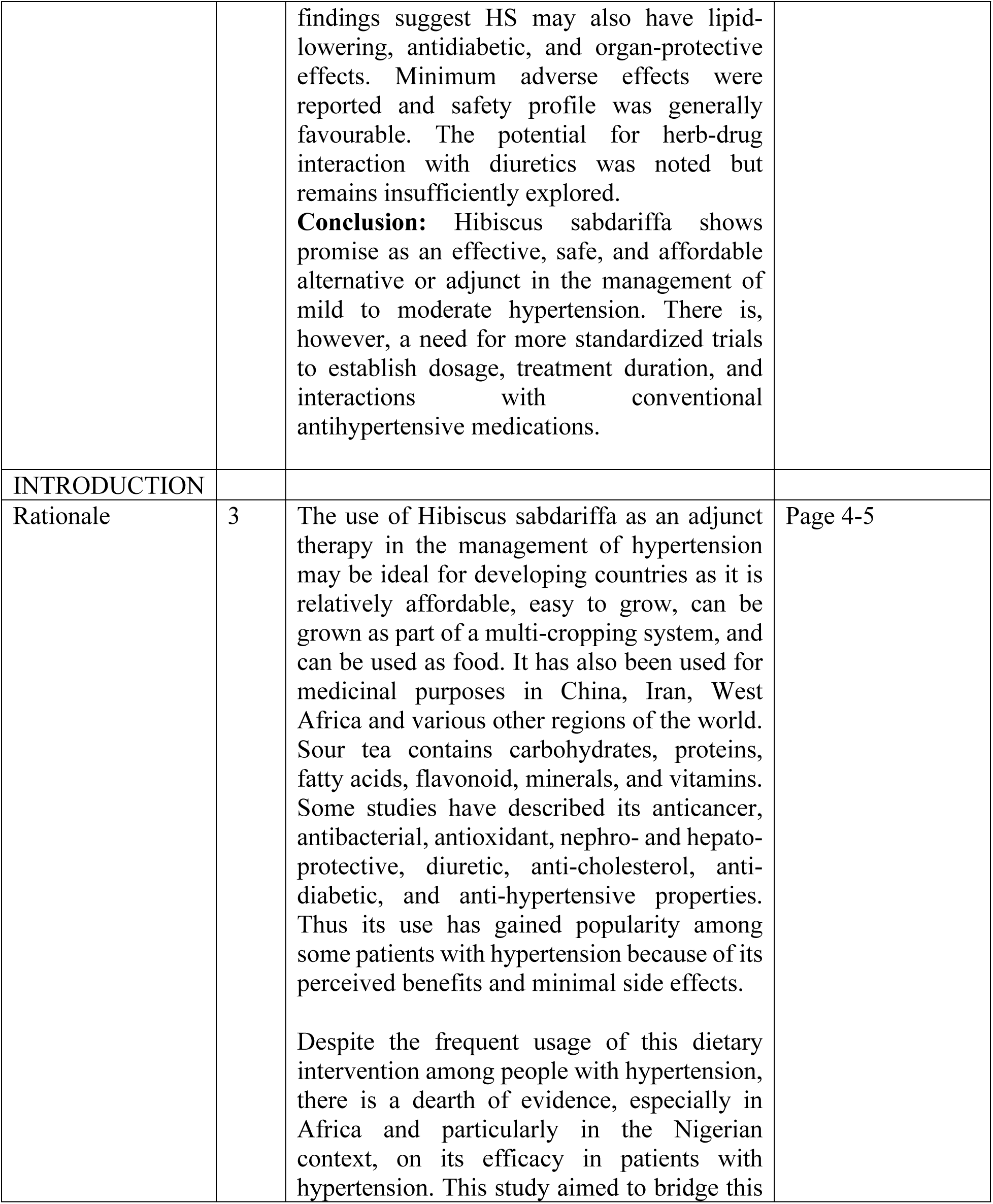
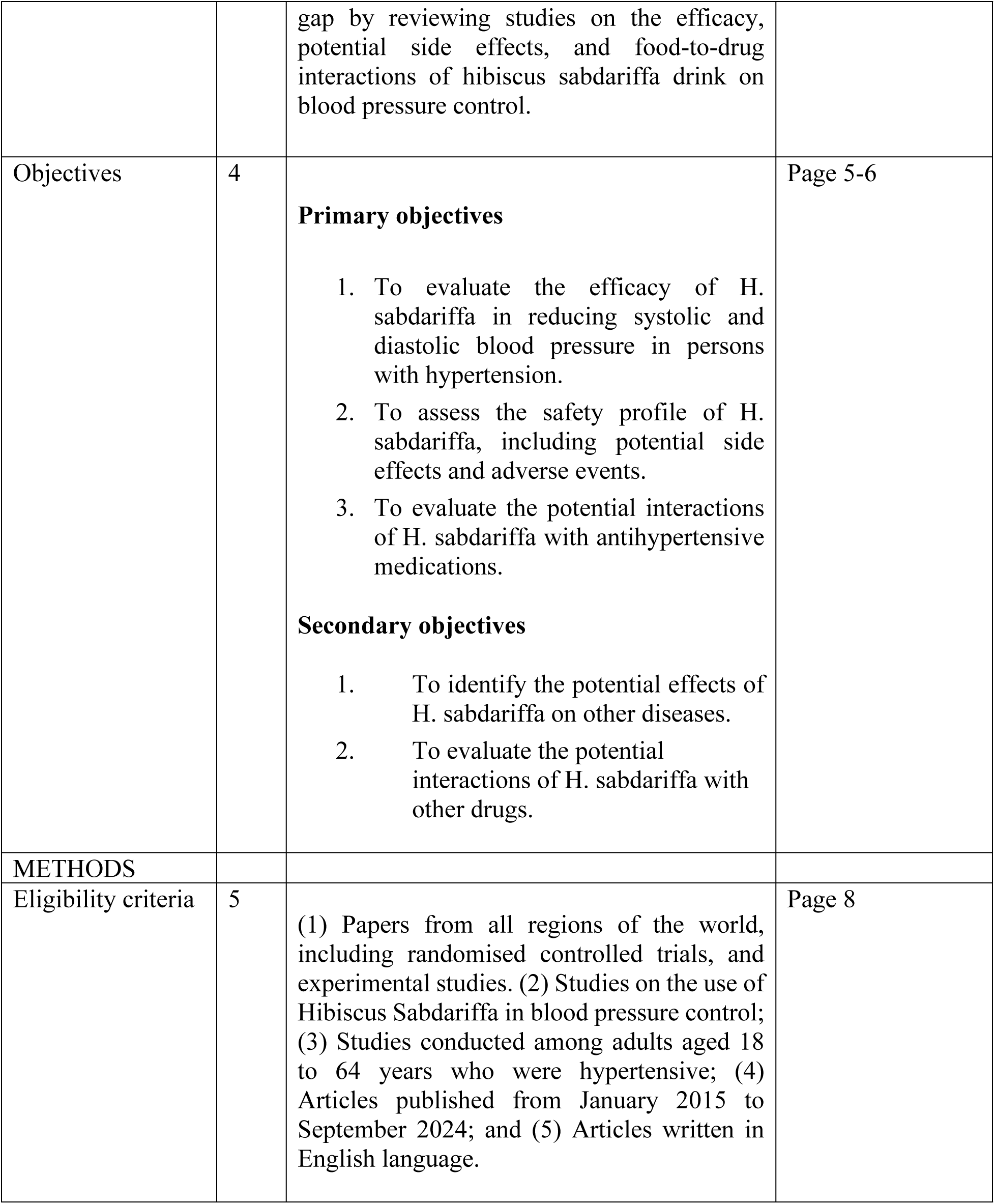
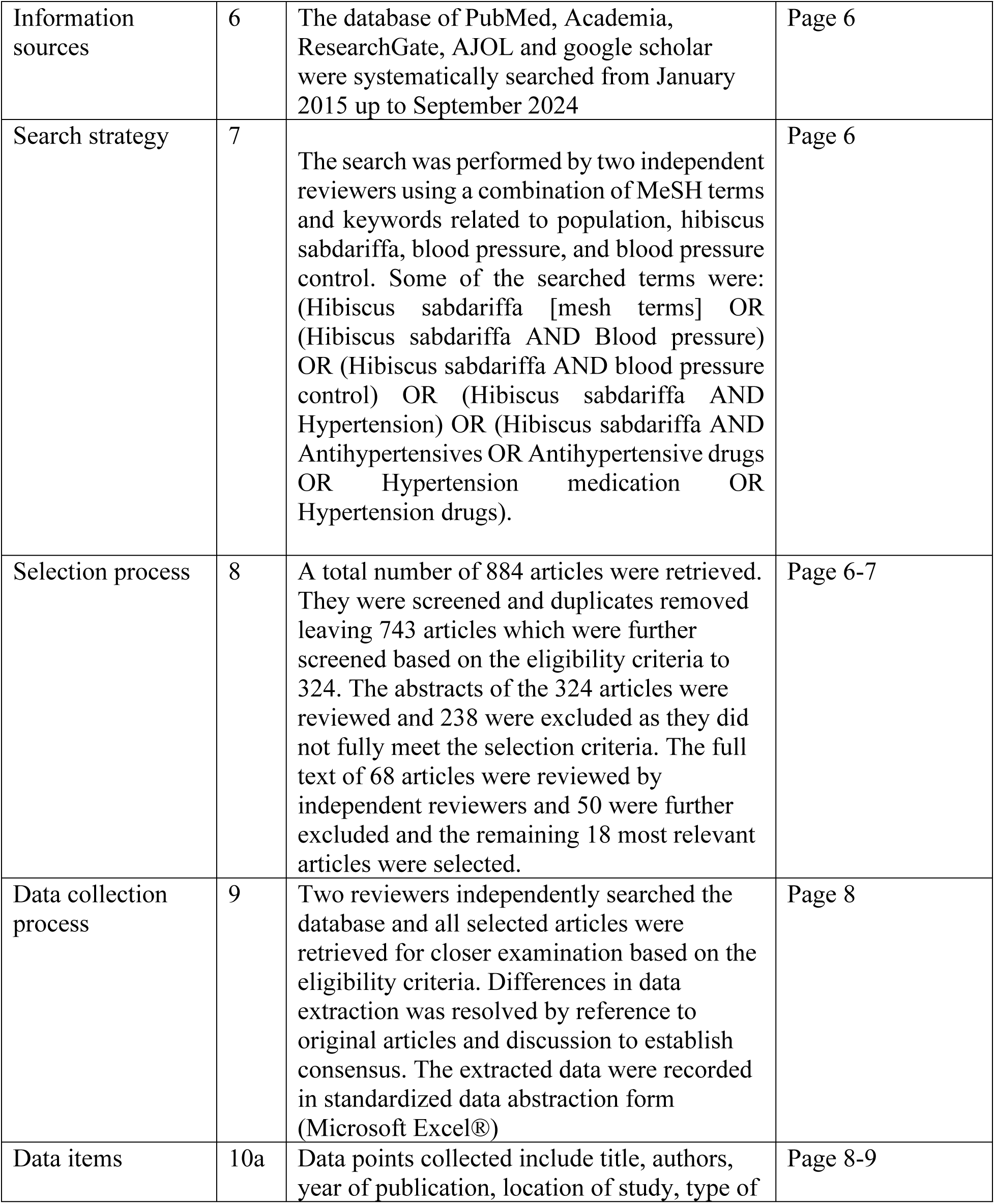
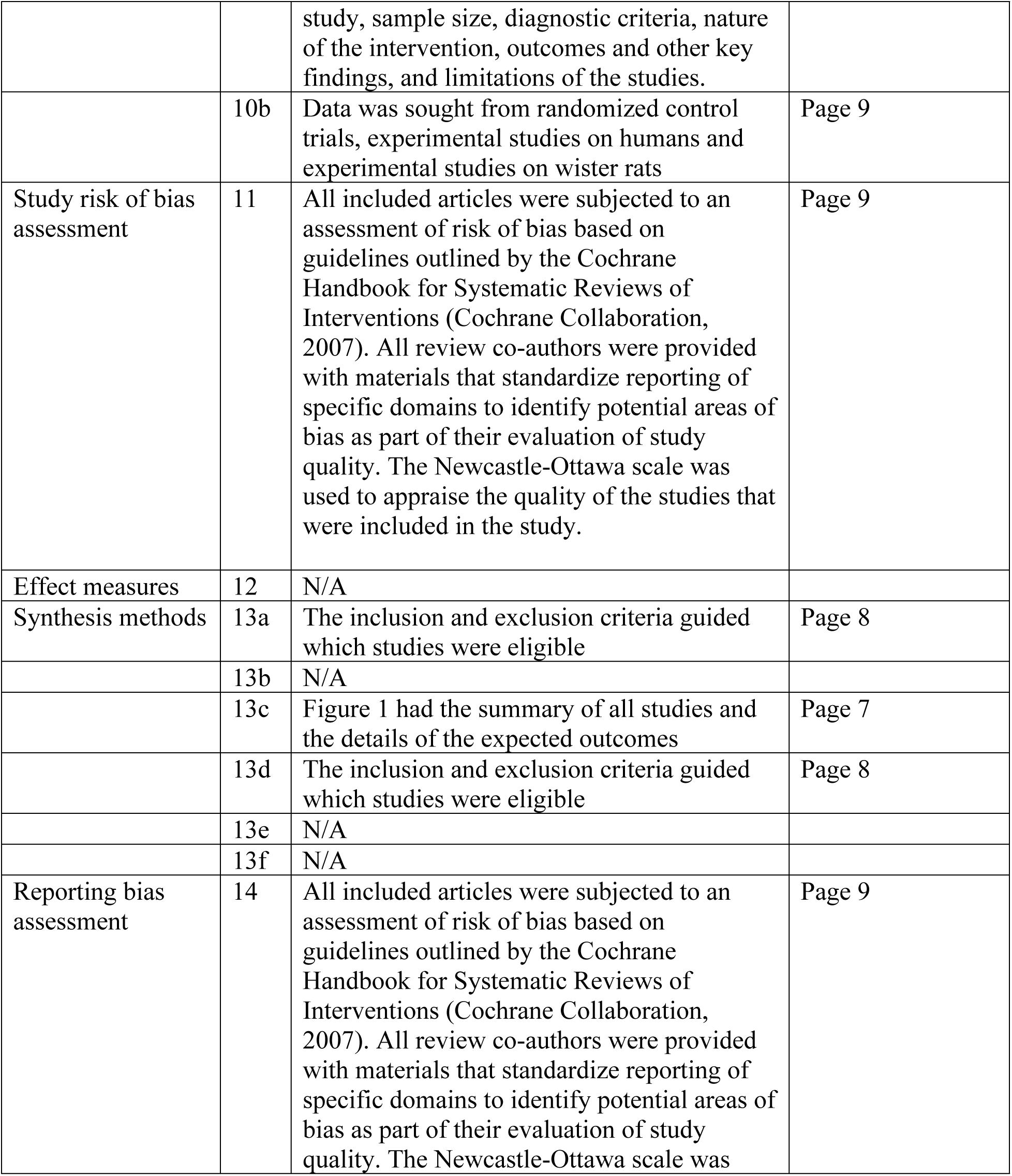
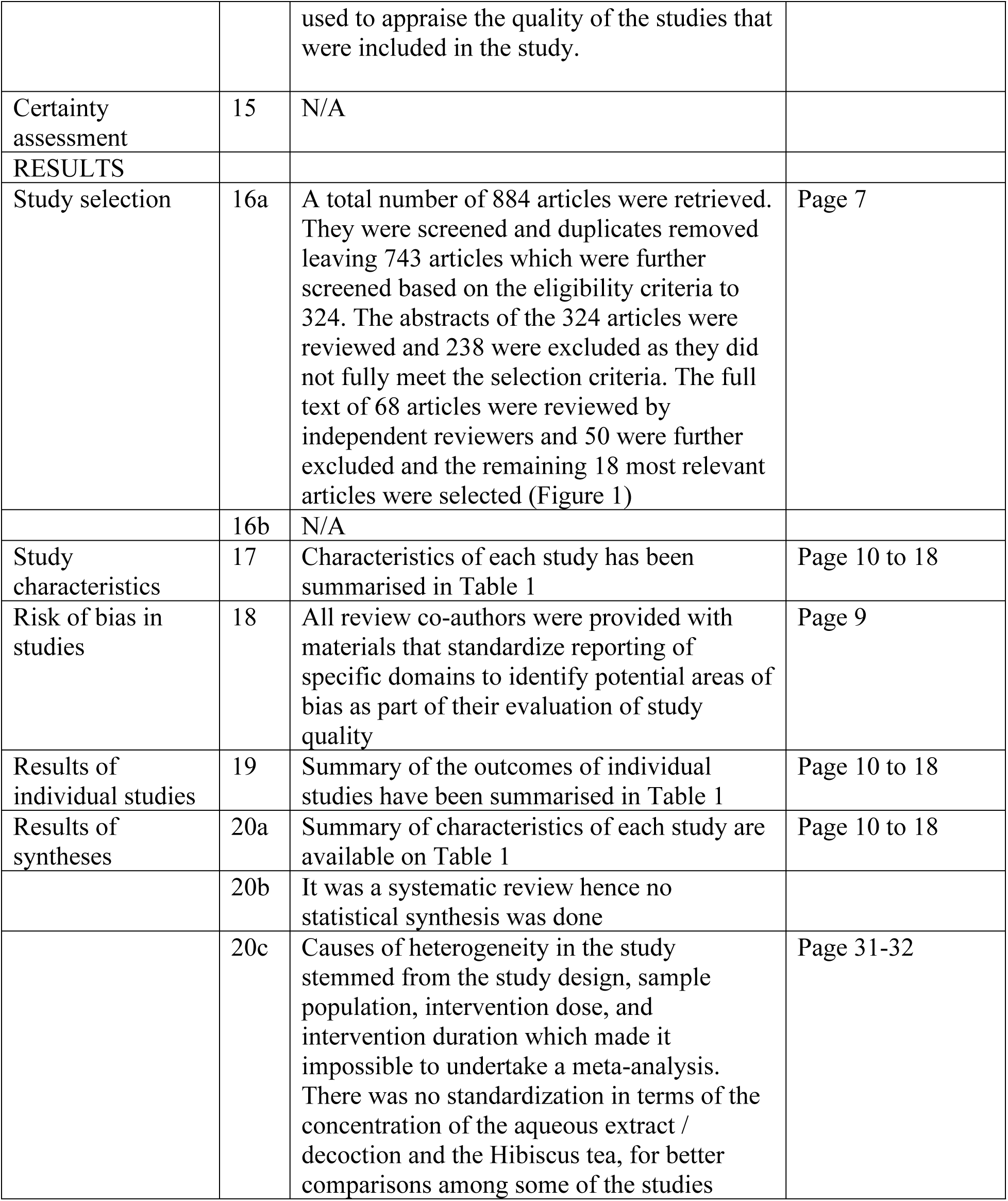
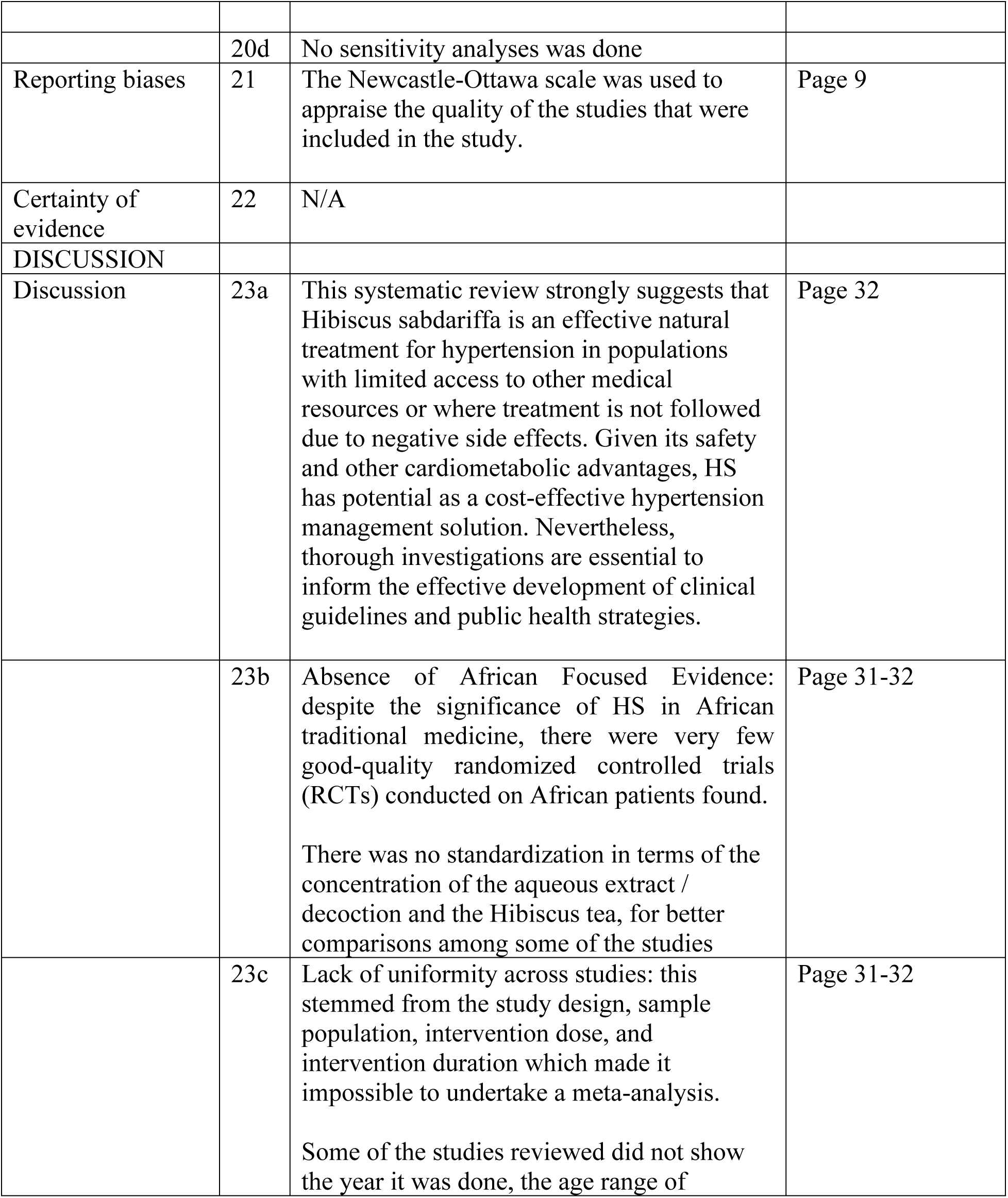
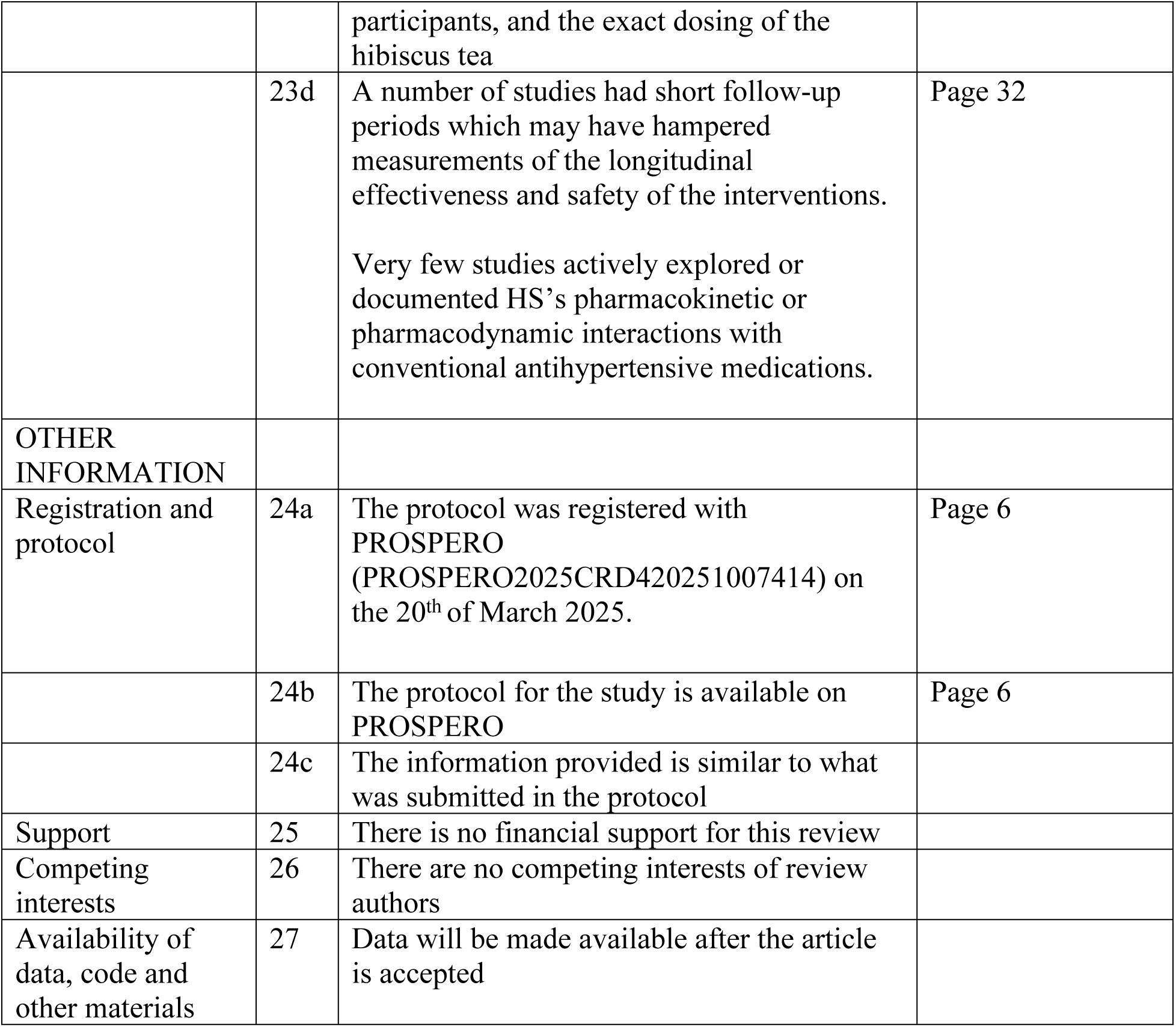

## Notes

### Competing Interest Statement

The authors have declared no competing interest.

### Funding Statement

The author(s) received no specific funding for this work.

### Author Declarations

Lagos State University Teaching Hospital HREC

